# Traces of SARS-CoV-2 RNA in the Blood of COVID-19 Patients

**DOI:** 10.1101/2020.05.10.20097055

**Authors:** Ahmed Moustafa, Ramy K. Aziz

**Affiliations:** Department of Biology, American University in Cairo, New Cairo, Egypt; Department of Microbiology and Immunology, Faculty of Pharmacy, Cairo University, Cairo, Egypt; Center for Genome and Microbiome Research, Cairo University, Cairo, Egypt

**Keywords:** SARS-CoV-2, Coronavirus, COVID-19, Blood, PBMC, RNA-Seq

## Abstract

The severe acute respiratory syndrome coronavirus 2 (SARS-CoV-2) is the third virus that caused coronavirus-related outbreaks over the past 20 years. The outbreak was first reported in December 2019 in Wuhan, China, but rapidly progressed into a pandemic of an unprecedented scale since the 1918 flu pandemic. Besides respiratory complications in COVID-19 patients, clinical characterizations of severe infection cases showed several other comorbidities, including multiple organ failure (liver, kidney, and heart) and septic shock. To better understand COVID-19 pathogenesis in different human organs, we interrogated the presence of the virus in the blood, or any of its components, which might provide a form of trafficking or hiding to the virus. By computationally analyzing high-throughput sequence data from patients with active COVID-19 infection, we found evidence of only traces of SARS-CoV-2 RNA in peripheral blood mononuclear cells (PBMC), while the virus RNA was abundant in bronchoalveolar lavage specimens from the same patients. We also devised a ‘viral spike-to-actin’ RNA normalization, as a metric to compare across various samples and minimize errors caused by intersample variability in human RNA. To the best of our knowledge, the presence of SARS-CoV-2 RNA in the PBMC of COVID-19 patients has not been reported before, and this observation could suggest immune presentation, but discounts the possibility of extensive viral infection of lymphocytes or monocytes.

## Introduction

The severe acute respiratory syndrome coronavirus 2 (SARS-CoV-2) is the third virus that caused coronavirus-related outbreaks over the past 20 years. The first outbreak occurred in Asia in 2002-2003 causing Severe Acute Respiratory Syndrome (SARS), hence, the name, SARS-CoV, which back then was not related to any of the known viruses (Marra et al. 2003; Rota et al. 2003). Between 2002 and 2003, 8,098 people became sick with SARS, and of those 774 died (i.e., a mortality rate of 9.5%). Since 2004, there have been no more reports of SARS cases (via [NHS](https://www.nhs.uk/conditions/sars/), [WHO](https://www.cdc.gov/sars/about/faq.html), and [CDC](https://www.cdc.gov/sars/about/faq.html)).

The second coronavirus-related outbreak started in the Arabian Peninsula in 2012 (Zaki et al. 2012) causing a more fatal disease, Middle East respiratory syndrome (MERS), with a significantly higher mortality rate of 40% of the cases infected by MERS-CoV virus (Zumla, Hui, and Perlman 2015).

More recently, in December 2019, the third coronavirus-related outbreak was first reported in Wuhan, China, by the emergence of the Severe Acute Respiratory Syndrome Coronavirus 2 (SARS-CoV-2), initially dubbed “the 2019 novel coronavirus" (2019-nCoV). The spread of the virus led to a pandemic of an unprecedented scale since the 1918 flu pandemic.

As of May 30, nearly 6 million confirmed COVID-19 cases globally and > 350,000 deaths have been reported by WHO ([WHO Dashboard](https://covid19.who.int/), continuously updated). Besides the respiratory complications in COVID-19 patients, clinical characterizations of severe infection cases indicated further comorbidities, including multiple organ failure (liver, kidney, and heart) and septic shock (Poston, Patel, and Davis 2020; Cascella et al. 2020; Hui Li et al. 2020).

The genome sequence of SARS-COV-2 has been determined and made public (Lu et al. 2020), and since then thousands (over 35,000) of genomes have been sequenced from all around the world (Shu and McCauley 2017)https://www.gisaid.org(Shu and McCauley 2017). The availability of those genomic sequences allows the rapid screening of viral RNA in human tissues as well as environmental samples (e.g., sewage (Bibby and Peccia 2013)) using multi-omic wet lab technologies, and *in silico* screening tools, for publicly available metatranscriptomic samples.

To better understand COVID-19 pathogenesis in different human organs, we conducted this study to interrogate the presence of the virus in the blood, or any of its components, as it might provide a form of trafficking or hiding to the virus, notably that some precarious studies reported the ability of the virus to infect lymphocytes (X. Wang et al. 2020) whereas others suggested the virus exerts its pathogenesis through “attacking hemoglobin,” although this hypothesis has been heavily criticized (Read 2020). The virus was reported to be found in the plasma of COVID-19 patients (Huang et al. 2020). Finally, peripheral blood mononuclear cells (PBMC) were shown to harbor other infectious viruses, such as HIV, HCV, and HBV (W.-K. Wang et al. 2002; Z. Li, Hou, and Cao 2015).

For the aforementioned reasons, we computationally analyzed high-throughput sequence data from patients with active COVID-19 infection, and found evidence of traces of SARS-CoV-2 RNA in their PMBCs, while their bronchoalveolar lavage samples had large amounts of the viral RNA.

## Methods

### Dataset

Publicly accessible raw RNA-Seq FASTQ sequences published by Xiong et al. (Xiong et al. 2020) were obtained from the Genome Sequence Archive (Y. Wang et al. 2017) (GSA accession CRA002390). Raw sequences were processed for quality control using fastp (Chen et al. 2018).

### Comparison of RNA Abundance and Gene Expression Profiles

Filtered FASTQ sequences were searched with blastx (Altschul et al. 1990) against the RefSeq protein database (O’Leary et al. 2016) (release 99) using DIAMOND (Buchfink, Xie, and Huson 2015) with an e-value cutoff < 1e-10. The counts of the matching RefSeq accessions were normalized to the total number of reads per library. Principal Component Analysis (PCA) was performed on the normalized abundance of the RefSeq proteins. For functional annotation, the matching RefSeq proteins were annotated by Pfam (Finn et al. 2014) using HMMER (Eddy 2011).

### Detection of Viral RNA

Filtered FASTQ sequences were aligned to the SARS-CoV-2 reference genome (GenBank accession [NC_045512](https://www.ncbi.nlm.nih.gov/nuccore/1798174254)) using the Burrows-Wheeler aligner (BWA, (Heng Li and Durbin 2009)). Generated binary alignment map (BAM) files were filtered for mapped sequences with a quality score > 40 and alignment score > 90 using Sambamba (Tarasov et al. 2015). Identified SARS-CoV-2 matching sequences were manually inspected and searched against NCBI “nt” by BLAST (blastn) for verification (Altschul et al. 1990).

### Data normalization

To estimate viral RNA abundance in a given sample, we normalized any positive hits to SARS-CoV-2 to the total number of reads within that sample. Additionally, we used the spike gene, being a highly specific SARS-CoV-2 gene to estimate the extent of viral RNA load in a given sample, and we normalized the abundance of spike genes to human actin RNA, being a transcript of a housekeeping gene. This normalization generated a ‘spike-to-actin’ ratio that could be used as an accurate metric for viral RNA abundance relative to human RNA.

## Results

We analyzed the RNA-Seq dataset published by Xiong et al. (Xiong et al. 2020). **Table S1** describes the processed samples, and the sequence reads statistics of each sample:

Based on the global gene expression in PBMC, PCA showed a strong, expected separation between BALF and PBMC samples, and further slight separation in the PBMC from healthy controls and COVID-19 patients (**Figure 1**). However, one healthy control was placed within the COVID-19 cluster.

**Figure 1.**
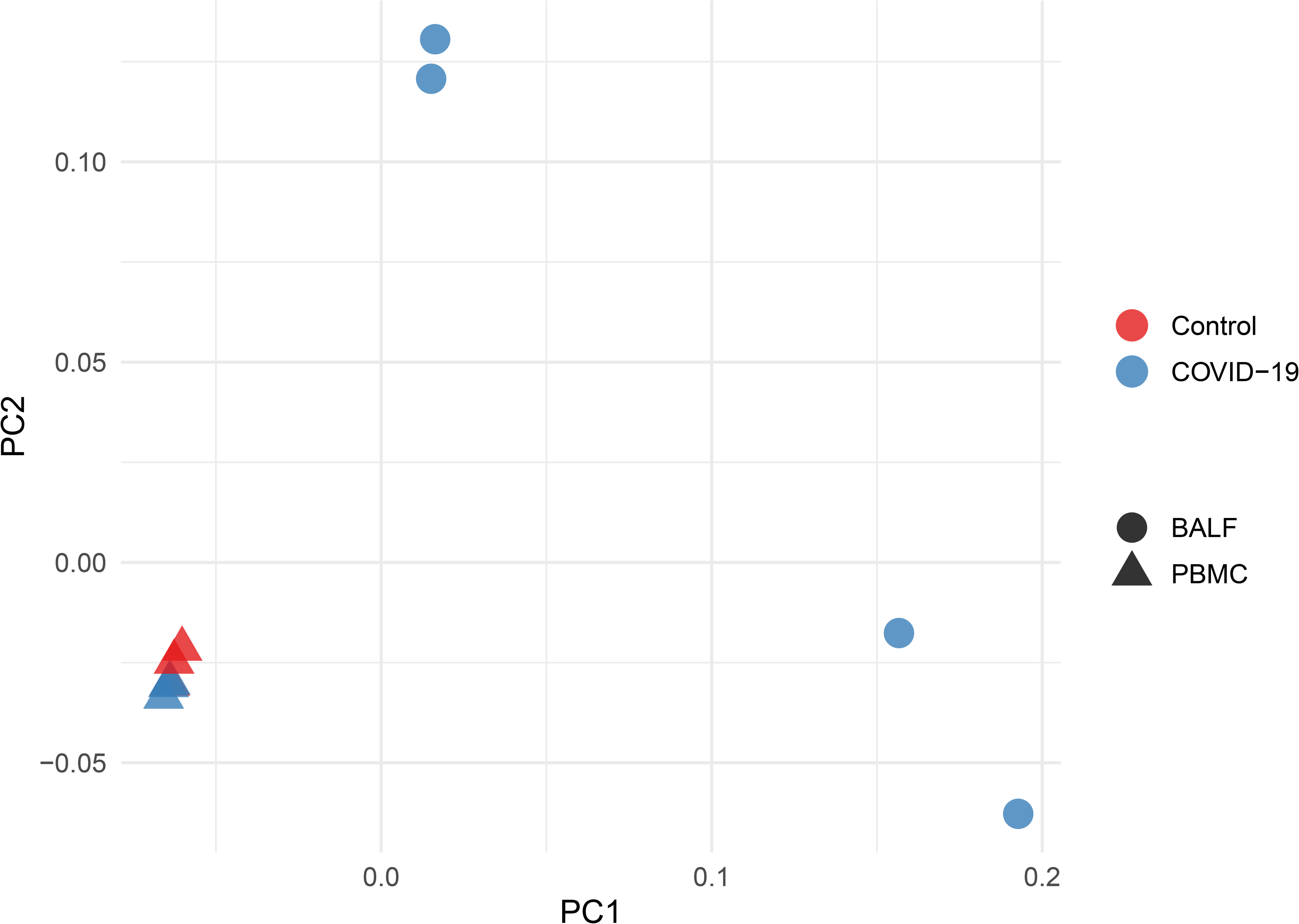
Principal Component Analysis based on RNA abundance data. Samples are color-coded; red for healthy donors, and green for COVID-19 patients. PC1 explains 63% of the variance and PC2 explains 30% of the variance.

We found viral sequences in all the BALF samples with a median of abundance 2.15% of the total reads (**Table S1**). We also identified 2 paired-ended viral reads in a PBMC sample of a COVID-19 patient, matching the SARS-CoV-2 polyprotein (pp1ab) (accession [NP_828849](https://www.ncbi.nlm.nih.gov/protein/30124074)) and SARS-CoV-2 surface glycoprotein (accession [YP_009724390](https://www.ncbi.nlm.nih.gov/protein/1796318598)) (**Figures 2, 3**).

**Figure 2.**
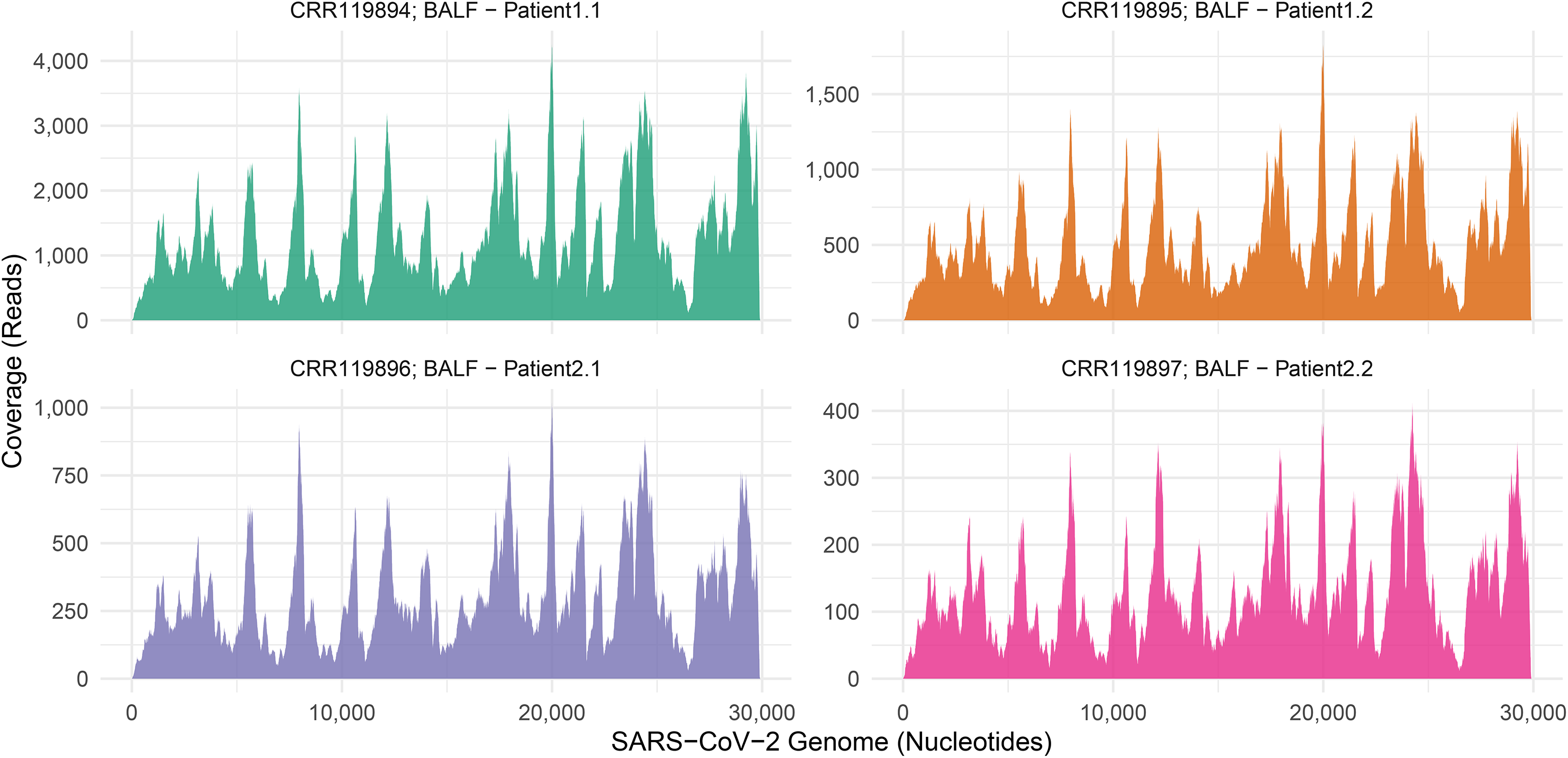
Mapping of SARS-CoV-2 RNA against the SARS-CoV-2 genome. Viral RNA sequences from the COVID-19 patients were mapped against the SARS-CoV-2 reference genome (GenBank accession [NC_045512](https://www.ncbi.nlm.nih.gov/nuccore/1798174254)). The x-axis is the nucleotide position on the virus genome. The y-axis is the coverage (in reads; not normalized) of the genomic position by RNA-Seq viral sequences. Panel “A” represents the BALF samples. Panel “B” represents the PBMC samples.

**Figure 3.**
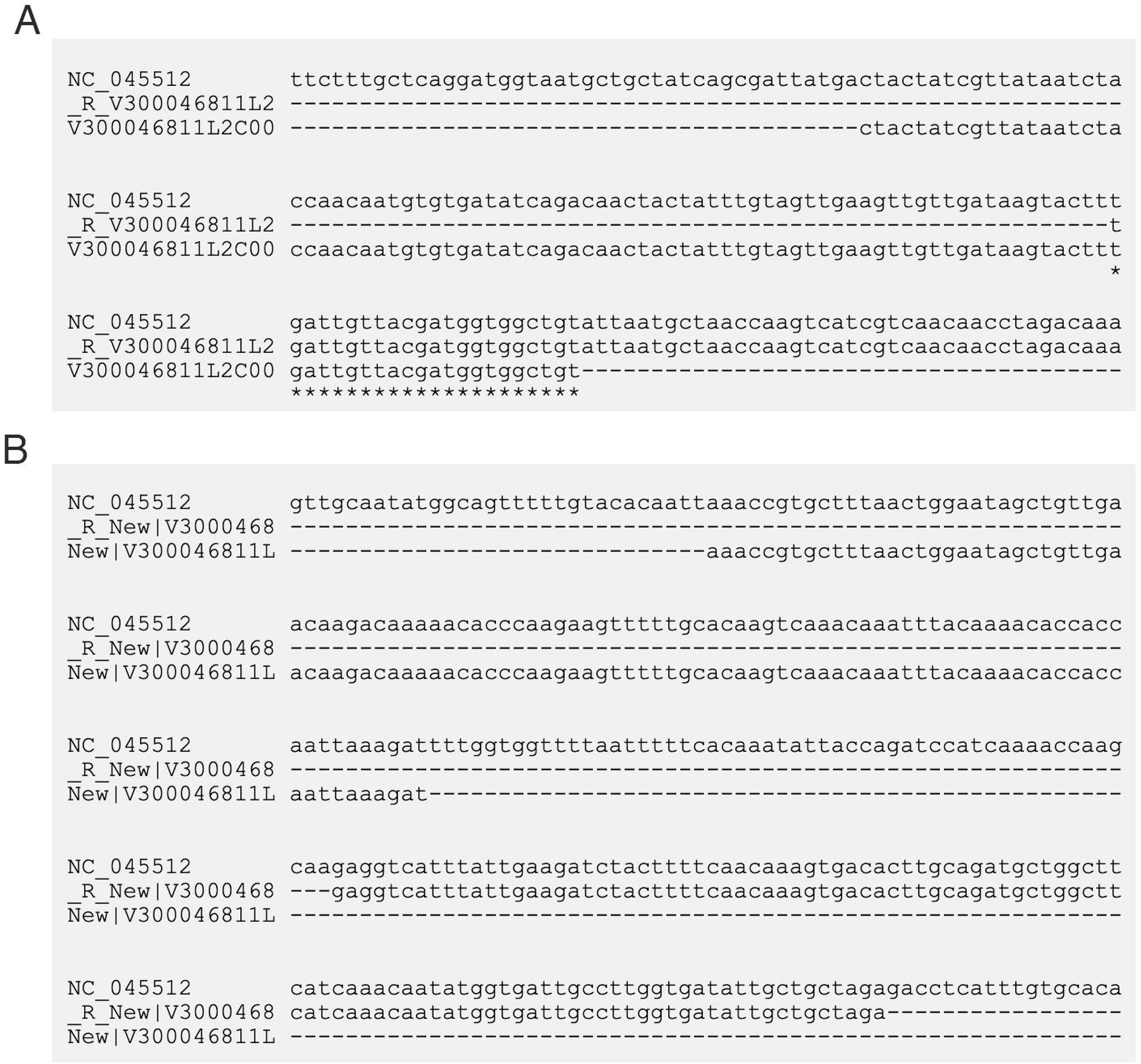
Alignment of the detected viral RNA sequences to the SARS-CoV-2 genome. The two paired-end viral reads from a COVID-19 PBMC sample aligned to the SARS-CoV-2 reference genome (GenBank accession [NC_045512](https://www.ncbi.nlm.nih.gov/nuccore/1798174254)). Panel A is the alignment first paired-end read. Panel B is the alignment second paired-end read. Perfect identities between the reference and the two overlapping mates are indicated by asterisks (“*”).

When we calculated a spike-to-actin RNA ratio for each sample, values for all four BALF samples, as well as the one PBMC sample with SARS-CoV-2 transcripts, followed the same pattern of viral RNA abundance values, and strongly correlated (Spearman correlation *p =* 0.96, *p*-value = 0.00047, **Figure 4C**)

**Figure 4.**
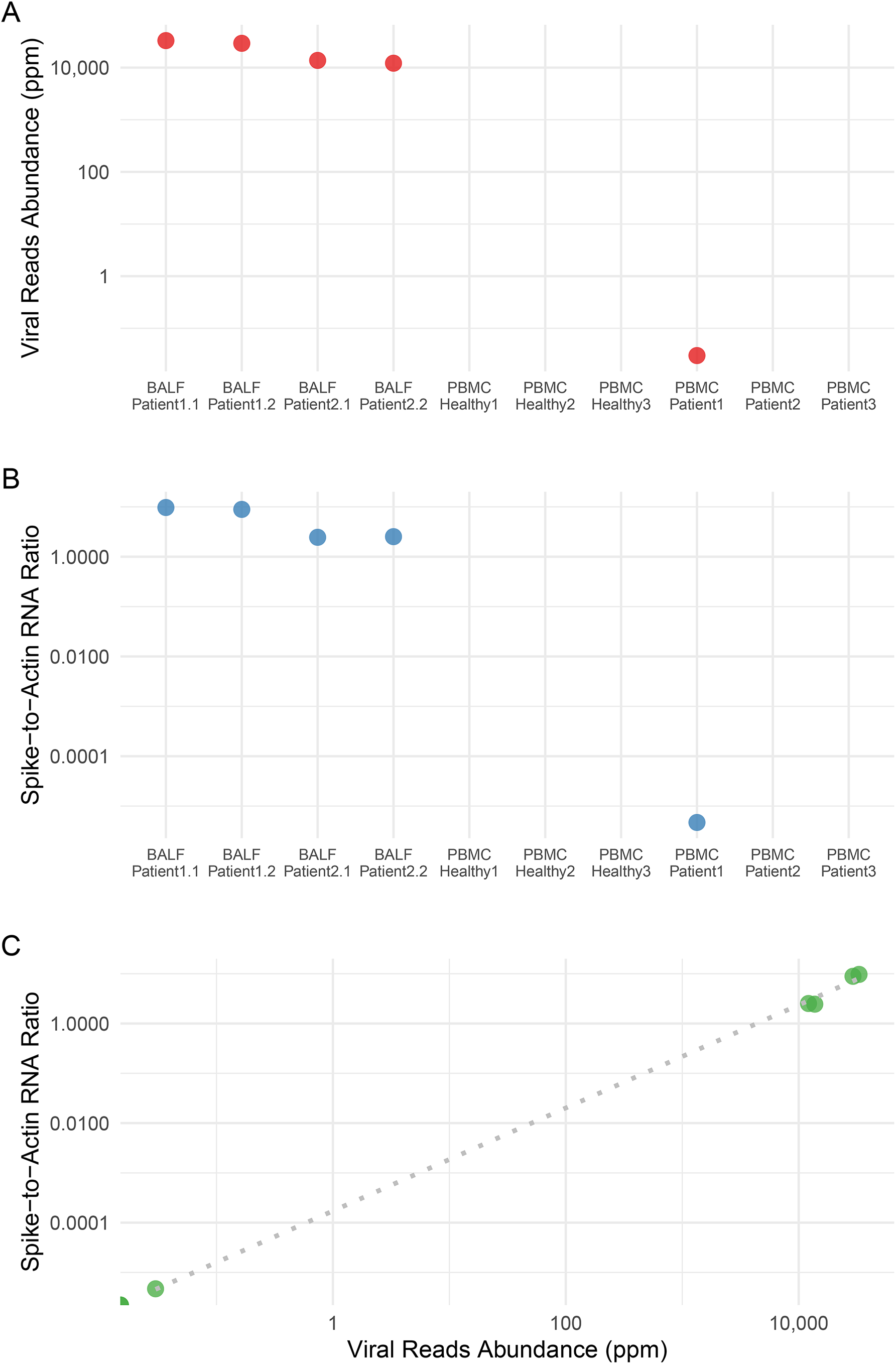
SARS-CoV-2 viral abundance relative to the host gene expression. A) Normalized abundance of SARS-CoV-2 viral reads, calculated number of SARS-CoV-2 specific hits per million sequence reads. B) SARS-CoV-2 spike-to-human actin ratio for each sample. C) A scatter plot showing the correlation between SARS-CoV-2 spike-to-human actin ratio and SARS-CoV-2 viral reads abundance (in ppm).

## Discussion

Coronavirus-related infections have been reported to be associated with hematological changes including lymphopenia, thrombocytopenia, and leukopenia through infecting blood cells, bone marrow stromal cells, or inducing autoantibodies (Yang et al. 2003).

In a former study characterizing the clinical features of COVID-19 patients, Huang and coworkers showed that using RT-PCR allowed them to detect coronavirus in plasma isolated samples from the patients (Huang et al. 2020). In their report, they preferred to use the term “RNAaemia,” rather than “viraemia,” which they defined as the presence of a virus in the blood because they did not perform tests to confirm the presence of an infectious SARS-CoV-2 virus in the blood of the patients.

There have been no reports of detecting viral RNA in blood cells, notably PBMC. In their pcreliminary analysis of RNA isolated from PBMC, [Corley et al](https://doi.org/10.1101/2020.04.13.039263) confirmed that they did not detect viral sequences (a preprint 10.1101/2020.04.13.039263v1).

Using high-throughput sequencing has been repeatedly demonstrated to be an effective approach for the identification and quantification of viruses in the blood (Moustafa et al. 2017) following similar methods as those used in viral metagenomics (Breitbart et al. 2003);(Aziz et al. 2015)) and uncultivated viral genomics ((Roux et al. 2019).

Therefore, we planned to exhaustively look for SARS-CoV-2 RNA sequences in publicly available RNA-Seq PBMC datasets. As of the writing of this report, there has been only one publicly available RNA-Seq PBMC dataset published by (Xiong et al. 2020), in which the group profiled global gene expression in BALF and PBMC specimens of COVID-19. Predictably, we detected viral RNA in all BALF samples (2 patients, 2 replicates each) with an average abundance of 2.15% of the total RNA in those samples, which included human RNA (**Table S1 and Figure 4**).

Of note, to estimate the viral RNA load in a given sample and to compare between different samples, we normalized viral read counts to the total number of reads in each data set. Since the total number of human-related reads may be strongly affected by potential microbial RNA (from pulmonary microbiota, for example) or by extensive viral counts, we also sought to estimate the ratio of viral genomic RNA to human cellular RNA. For that purpose, we used actin RNA, being a transcript for a human housekeeping gene—not expected to be seen in bacterial cells or viral genomes, and we estimated the ratio of hits to RNA encoding the spike protein (S) copies to actin transcripts in the sample. Quite interestingly, the spike-to-actin ratio strongly correlated with viral RNA abundance values (computed as SARS-CoV-2 RNA reads normalized to the total number of reads), **Figure 4C**.

As for blood samples, we identified two paired-end reads in RNA isolated from PBMC from only one (accession CRR119891) out of three patients, which aligned to the SARS-CoV-2 genome. Expectedly, no SARS-CoV-2 RNA was detected in the PBMC from healthy controls (three donors).

These RNA traces are certainly quite rare; however, they confidently and specifically belong to SARS-COV-2 (**Figure 3**). One viral RNA read translates into polyprotein (pp1ab, accession [NP_828849](https://www.ncbi.nlm.nih.gov/protein/30124074)), which is the largest protein of coronaviruses and involved in the replication and transcription of the viral genome. The other viral RNA translates into surface (spike) glycoprotein (accession [YP_009724390](https://www.ncbi.nlm.nih.gov/protein/1796318598)), which mediates the entry of the viral into the human cells expressing human angiotensin-converting enzyme 2 (hACE2) (Ou et al. 2020; Walls et al. 2020). Although we are not rejecting the possibility of cross-contamination or barcode bleeding (Mitra et al. 2015; Kircher, Sawyer, and Meyer 2012) for detecting the viral RNA in one PBMC RNA-Seq sample, such possibility is unlikely, given that control samples had zero hits to SARS-CoV-2 RNA. We are also considering the possibility of SARS-CoV-2 being sampled by antigen-presenting cells (most likely dendritic cells) or presented to T lymphocytes, which are in the PBMC population.

One more possibility, which we believe needs many more samples to consider, is that SARS-CoV-2 may be specifically or coincidentally internalized by one of the mononuclear cell types, which may suggest a mechanism for the chronicity of the SARS-CoV-2 infection. This hypothesis requires further testing. However, in the light of our data, it is hard to support the early reports that SARS-COV-2 targets T-lymphocytes *in vivo*, as suggested earlier, in a correspondence, based on cell culture experiment with pseudotyped viruses (X. Wang et al. 2020).

With more data becoming publicly available, it will be possible to revisit this hypothesis and others to improve our understanding of the progression and replication of SARS-CoV-2 in infected individuals.

## Data Availability

The data was published by Xiong et al. 2020 and is publicly available through the Genome Sequence Archive with accession CRA002390.

https://bigd.big.ac.cn/gsa/browse/CRA002390

